# Apathy progression is associated with brain atrophy and white matter damage in Parkinson’s disease

**DOI:** 10.1101/2025.01.22.25320946

**Authors:** Roqaie Moqadam, Houman Azizi, Aliza Brzezinski-Rittner, Lucas Ronat, Alexandru Hanganu, Yashar Zeighami, Mahsa Dadar

**Affiliations:** Department of Medicine, University of Montreal, Montreal, Quebec, Canada; Douglas Mental Health University Institute, Montreal, Quebec, Canada; Centre de Recherche, Institut Universitaire de Gériatrie de Montréal, Québec, Canada; Montreal Neurological Institute and Hospital, McGill University, Montreal, Quebec, Canada; Integrated Program in Neuroscience, McGill University, Montreal, Quebec, Canada; Department of Psychology, University of Montreal, Montreal, Quebec, Canada; Department of Psychiatry, McGill University, Montreal, Quebec, Canada

**Author notes:** Correspondence to: Mahsa Dadar, Douglas Mental Health University Institute, Montreal, Quebec, Canada; Roqaie Moqadam, Douglas Mental Health University Institute, Montreal, Quebec, Canada. These authors contributed equally to this work (Yashar Zeighami, Mahsa Dadar).

**Keywords:** Parkinson’s disease, apathy, magnetic resonance imaging, deformation-based morphometry, white matter hyperintensities, longitudinal analysis

## Abstract

Apathy is a prevalent non-motor symptom that significantly impacts the quality of life in Parkinson’s disease (PD) patients. Although previous studies have investigated the neural correlates of apathy in PD, the longitudinal relationships between regional brain atrophy, white matter hyperintensities (WMHs), and apathy progression remain underexplored. Using longitudinal, multisite data of *de novo* PD patients from the Parkinson’s Progression Markers Initiative (PPMI), the present study aims to investigate these relationships.

We used T1-weighted magnetic resonance imaging (MRI) and clinical data from 445 participants. Apathy was assessed as part of the Movement Disorder Society Unified Parkinson’s Disease Rating Scale (MDS-UPDRS) Part I. We applied deformation-based morphometry (DBM) to quantify gray matter atrophy and used the Brain tISsue segmentatiON (BISON) algorithm to segment WMHs from T1-weighted images. Using linear regression models, we performed cross-sectional analyses to identify the associations between baseline brain measurements (DBM and WMH) and apathy severity. Longitudinal analyses utilized linear mixed-effects models to investigate whether baseline brain measurements were associated with future apathy progression over time, accounting for covariates such as age, sex, motion artifacts, Hoehn and Yahr stage, levodopa-equivalent daily dose (LEDD), Total Intracranial Volume (TIV), and baseline apathy. Hypothesis-based and exploratory analyses were conducted to confirm the results previously reported in the literature and explore potential new associations.

No cross-sectional regional associations survived multiple comparison corrections. Longitudinal hypothesis-based models confirmed that baseline atrophy in regions such as the bilateral nucleus accumbens area, superior parietal, putamen, insula, left precuneus, right precentral, and cerebellum gray matter was significantly associated with future apathy progression. Exploratory longitudinal analyses identified additional regions, including the bilateral lingual, parahippocampal, basal forebrain, ventral diencephalon, isthmus cingulate, thalamus, hippocampus, left middle temporal, right inferior temporal, pericalcarine, medial orbitofrontal, cuneus, where baseline atrophy was correlated with progression of apathy severity. Moreover, greater WMH burden, particularly in the frontal lobe, was associated with worsening apathy. These results highlight the influence of both gray matter atrophy and WMHs on apathy progression in PD.

## Introduction

Parkinson’s disease (PD) is the second most common neurodegenerative disorder after Alzheimer’s disease. In addition to its well-known motor symptoms including tremors, bradykinesia, rigidity, and postural instability, PD is associated with a range of non-motor symptoms such as apathy, depression, anxiety, cognitive impairment, sleep disturbances, and autonomic dysfunction^1,2^. Characterized by a lack of motivation, interest, and emotional engagement, apathy is one of the common neuropsychiatric symptoms in PD, substantially impacting the quality of life and independence of the patients^2–5^.

Stuss and colleagues define three distinct subtypes of apathy, including behavioral, motivational, and cognitive apathy, each associated with specific neural circuits^6–8^. While apathy is also present in up to 30% of healthy older adults, the apathy observed in PD patients is believed to differ from that associated with healthy aging^6,9^. This difference is primarily due to its association with specific neurotransmitter dysfunctions and alterations in apathetic PD patients^10^. As such, PD apathy is associated with dopaminergic deficits and disruptions in fronto-striatal circuits. These dysfunctions negatively impact reward processing, intrinsic motivation, and emotion regulation^6,10,11^. Structural changes in areas such as the basal ganglia and frontal cortex are thought to contribute to these deficits, emphasizing how PD-related apathy might differ from the aging-related alterations in motivation and engagement^6^.

The literature reports varying results in the prevalence of apathy across different PD populations, with a wide range from 13.9% to 70% depending on different factors including demographics, diagnostic criteria and assessment tools, disease stage, and treatment^3,4,12^. While apathy can emerge at any stage of the disease, it is frequently present in earlier disease stages^6^. In a recent review of the non-motor aspects of PD symptomatology, apathy was recognized as a distinct feature, occurring several years before the motor symptoms became evident^2^. In a cohort of 109 newly diagnosed untreated PD patients, apathy was commonly reported during the two-year period preceding the motor symptoms, with approximately 11% of the cohort identifying its onset within the period preceding the motor symptoms^13^.

Previous neuroimaging studies in cross-sectional samples have reported associations between cortical and subcortical atrophy and apathy in PD patients^14–23^. These studies vary in their approach to evaluating apathy, using either presence or severity measures. Regarding presence of apathy, several studies used the Neuropsychiatric Inventory (NPI)^24^ and Lille Apathy Rating Scale (LARS)^25^ to identify brain regions associated with apathy. These studies consistently reported associations between apathy presence and atrophy in regions such as the anterior cingulate cortex, nucleus accumbens, and caudate, emphasizing the involvement of frontostriatal circuits in presence of apathy^15–21^. Studies focusing on apathy severity often utilize the Starkstein Apathy Scale (SAS)^26^, Apathy Evaluation Scale (AES-I)^14,27^, and NPI. These studies established correlations between the severity of apathy and atrophy in certain brain regions including the inferior parietal lobule, orbitofrontal cortex, and insula, underscoring the relationship between cortical degeneration and apathy severity^14,15,19,20,22,23^ (Table 1). Longitudinal assessments of apathy related to brain atrophy have been more scarce. Morris et al^21^. reported an association between the nucleus accumbens volume and the presence of apathy over 2 years of follow-up. In contrast, Ranjbar et al^23^. reported a correlation between lower atrophy in the right putamen and increased apathy severity over time.

**Table 1.**
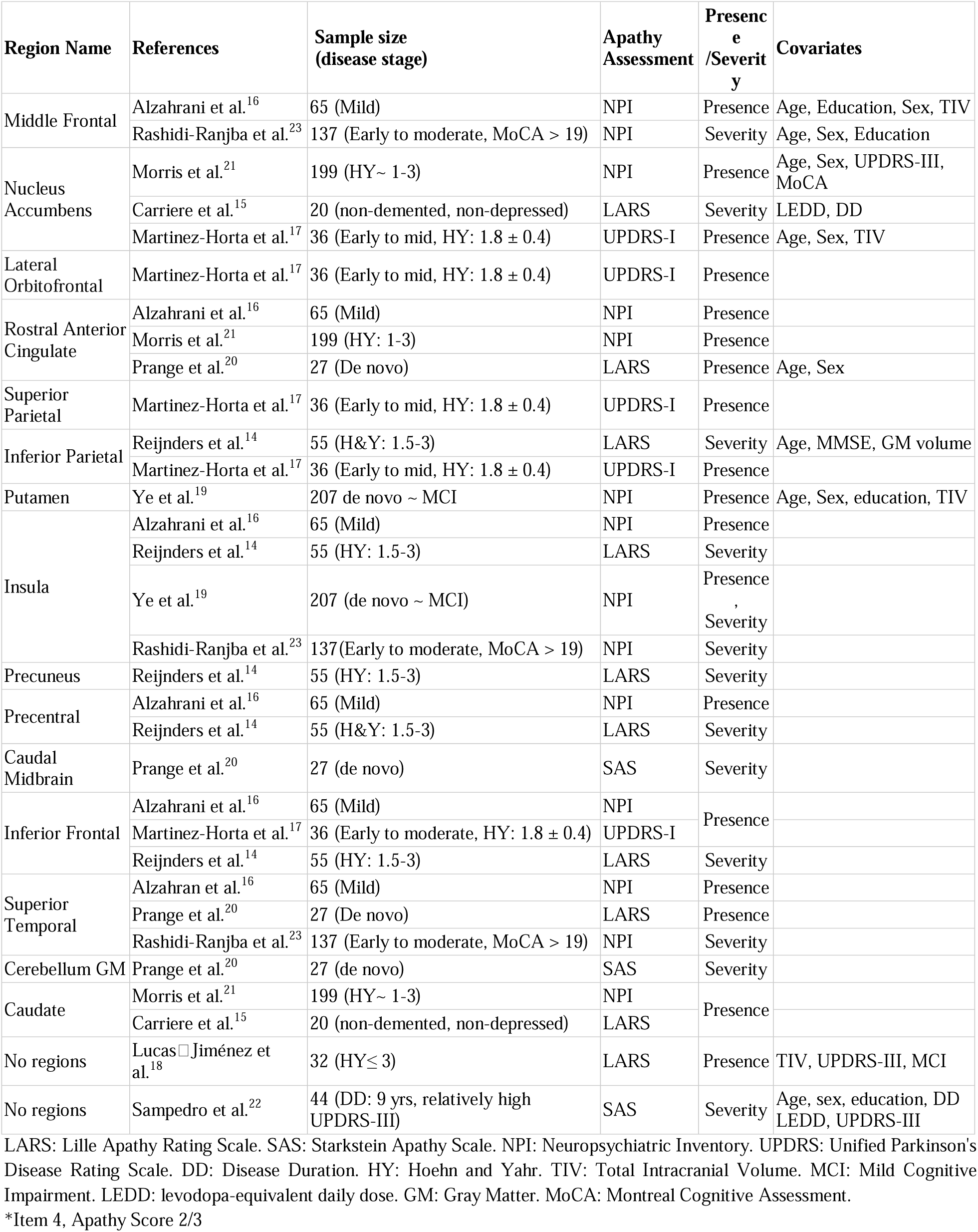
Regions linked to apathy in PD based on previous literature.

Beyond associations with gray matter atrophy, apathy has been linked to spatially extensive reductions in white matter (WM) integrity^28^. Using diffusion-weighted imaging (DWI), Wen et al^29^. found a decrease in WM network efficiency and connectivity related to apathy in PD, particularly within the frontal, temporal, parietal, and basal ganglia regions. However, DWI is not frequently used clinically, particularly in the context of PD^30^. On the other hand, in normal aging and individuals with Alzheimer’s disease^28,31^, the literature reports significant associations between apathy symptoms and increased volume of white matter hyperintensities (WMHs) in frontal and temporal lobes, areas critical for motivation and emotional regulation^28^. WMHs are areas of increased signal on fluid-attenuated inversion recovery (FLAIR) sequences and are markers of cerebrovascular pathology^32^. WMHs are also commonly present in patients with PD^33^. The association between WMHs and apathy in PD remains understudied, with a single study in a cohort of 141 PD patients reporting a significant link between WMH and apathy, where WMH severity was a strong predictor of apathy severity and progression in the patients^30^. However, this study used visual Fazekas scale for WMH assessment^34^, which does not reflect the full extent of WMH burden. In addition, only global WMH rating was utilized and the study did not assess how WMHs in different brain regions contribute to apathy^30^.

Although neural correlates of apathy have previously been examined, longitudinal relationships between regional cerebral atrophy, WMHs, and apathy require further investigation. The present study takes advantage of the large sample size and the availability of longitudinal clinical data from the Parkinson’s Progression Markers Initiative (PPMI) dataset to examine the relationship between apathy, regional brain atrophy, and WMH in patients with PD. We hypothesize that regional brain atrophy and WMH are associated with the severity and future progression of apathy in PD patients.

## Materials and methods

### Data

T1-weighted MRI and clinical data from 546 participants with PD were obtained from the Parkinson’s Progression Markers Initiative (PPMI) database (www.ppmi-info.org/data)^35^. The PPMI is a longitudinal, multi-site observational study of newly diagnosed and untreated patients with PD across the United States and Europe. Clinical assessments included Hoehn and Yahr stage (HY Stage), and apathy scores based on the Movement Disorders Society Unified Parkinson’s Disease Rating Scale part I (MDS-UPDRS part I)^35,36^. Levodopa equivalent daily dosage (LEDD) was calculated based on the dosage and type of the dopaminergic medications taken at the time of the each visit according to the guidelines provided by the PPMI (https://www.ppmi-info.org/sites/default/files/docs/PPMI Data User Guide.pdf). Clinical and demographic data were organized and processed through our open source data wrangling pipeline (https://github.com/HoumanAzizi/PPMI_Data_Wrangling). Participants who had received medication at baseline, or had missing MRI, demographics, or clinical information were excluded from the present study.

### MRI preprocessing

Figure 1 provides a schematic of the MRI processing steps performed. T1-weighted MRIs were processed using our open-source image processing pipeline based on the open-access MINC-Toolkit v2 (https://bic-mni.github.io/) and Advanced Normalization Tools (ANTs) (https://stnava.github.io/ANTs/) toolkits (https://github.com/VANDAlab/Preprocessing_Pipeline). Preprocessing steps included noise reduction^37^, intensity non-uniformity correction^38^, and intensity normalization into the range [0– 100]. The preprocessed T1-weighted images were first linearly^39^ and then nonlinearly^40^ registered to the MNI-ICBM152 average template^41^. All the image processing steps (i.e. pre-processing, linear, and nonlinear registrations) were visually quality controlled by an experienced rater (R.M.) blind to the clinical diagnosis, and the cases that did not pass this quality control step (N = 44) were excluded from the subsequent analyses.

**Figure 1.**
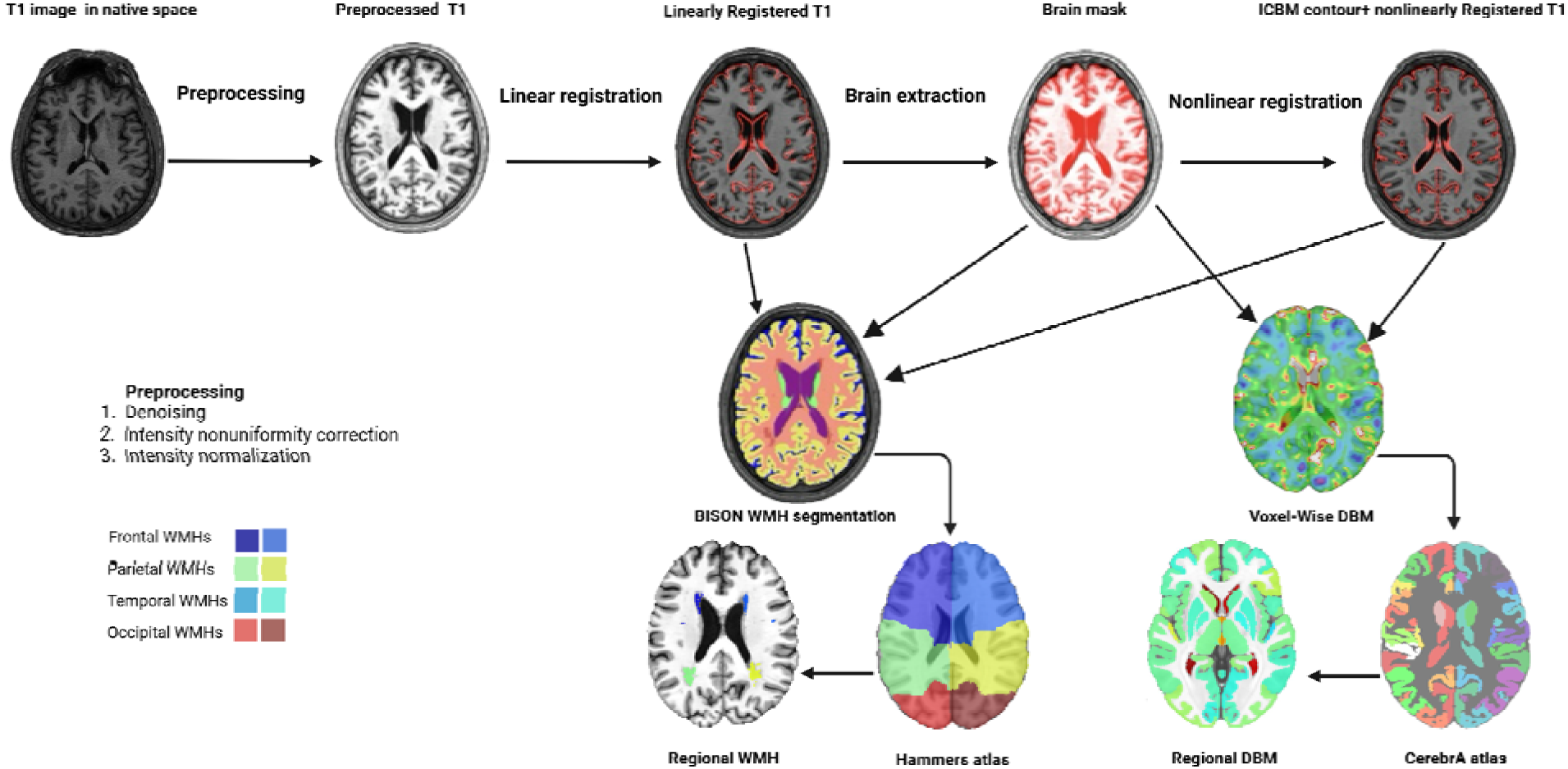
Flow diagram of the MRI processing steps performed. Native T1w images were preprocessed and linearly registered to the MNI-ICBM152 average template. Nonlinear registration was performed following brain extraction to derive voxel-wise DBM maps. CerebrA atlas was then used to extract regional DBM measures. The outputs of the linear registration, brain mask, and nonlinear registration steps were also used by BISON to derive voxel-wis WMH segmentations. Hammers lobar atlas was used to extract regional WMH measures. DBM: Deformation-base morphometry. WMH: White Matter Hyperintensity.

### Deformation-based morphometry (DBM)

Deformation-based morphometry (DBM) was used to extract regional gray matter atrophy, calculated as the Jacobian determinant of the deformation field from the nonlinear transformations^42^. DBM values reflect the relative volume of each voxel compared to the MNI-ICBM152 template, i.e. a value of 1 denotes the same volume compared to its equivalent voxel in the template, a value below 1 denotes a smaller volume than the corresponding voxel in the template, and a value above 1 denotes larger volume than the corresponding voxel in the template. Therefore, atrophy for a particular region can be inferred from decrease in DBM values. Average gray matter atrophy in 102 cortical and subcortical regions were determined using the CerebrA atlas^41^.

### WMH measurements

BISON^43^, a previously validated automatic segmentation tool was used to segment the WMHs. BISON combines a Random Forest classifier with a collection of location and intensity features obtained from a library of manually segmented scans to detect WMHs (Figure 1). Since the PPMI dataset lacked consistently acquired T2-weighted and Fluid attenuated inversion recovery (FLAIR) images for all participants, T1-weighted images were utilized for WMH segmentation. Although FLAIR and T2-weighted images are ideal for performing WMH assessments, in our previous work, we have shown that T1w-based segmentations from BISON yield estimates with strong correlations with FLAIR-based segmentations (r = 0.96)^44^, and have been previously used to assess WMH burden based on T1w images in multi-center cohorts, including the PPMI^45–47^. The WMH segmentations were visually quality controlled by an experienced rater (R.M.), and cases that did not pass this quality control step (N = 7) were excluded from the analyses. The number of voxels designated as WMH (measured in mm^3^) in the standard space (i.e. adjusted for intracranial volume) in each brain lobe and hemisphere (based on Hammer’s lobar atlas^44,48^) a well as across the entire brain were used to define regional and global WMH volumes, respectively^44,49^. WMH volumes were log-transformed to obtain a normal distribution.

### Motion assessment

Previous research including ours has shown a subtle but consistent impact of motion on gray matter atrophy estimations ^50–52^. As such, all the images that passed the QC steps outlined in the previous steps further underwent a visual inspection process to assess the presence and severity of motion similar to our previous work (R.M.)^52^. Based on our previous findings, cases with motion severity ratings greater than 3 (N = 27) were excluded^52^. Motion severity scores were further used as covariates in all models^52^(Figure 2).

**Figure 2.**
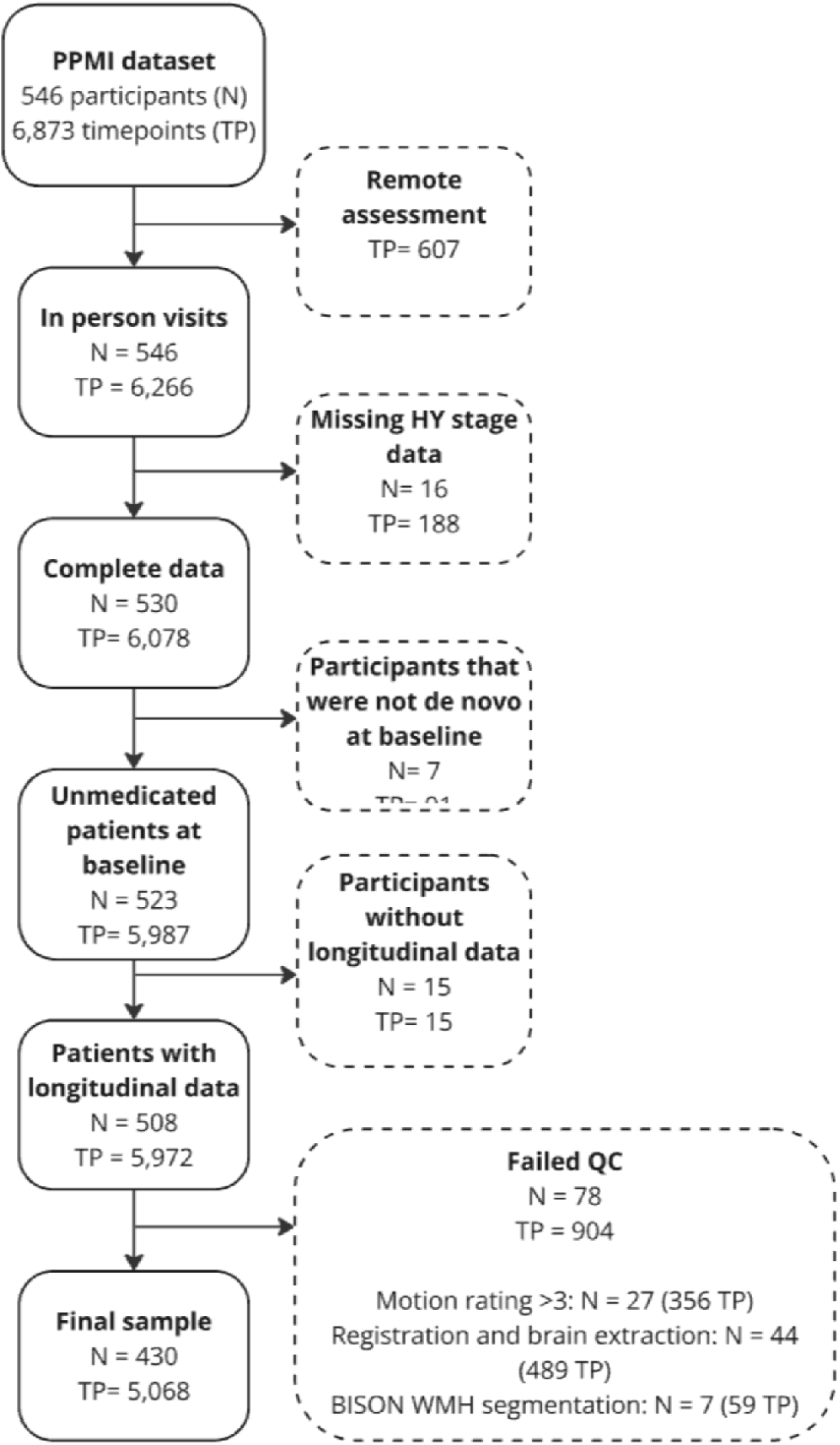
Diagram summarizing the participant selection procedure. A total of 116 participants were excluded due to unavailability of clinical assessments, presence of MRI artifacts, or failure of image processing steps, leading to a final sample of 430 de novo PD patients with 5,068 longitudinal clinical assessments. TP: Time point. QC: Quality control. HY: Hoehn and Yahr.

### Apathy assessment

Apathy was assessed using the Unified Parkinson’s Disease Rating Scale (UPDRS) part I with the following criteria: 0 represents no apathy, 1 indicates slight apathy, when the patient experiences apathy less than one day per week, 2 denotes mild apathy, when the patient experiences apathy more than one day per week but less than half of the days, 3 indicates moderate apathy, when the patient feels apathetic more than half of the days, 4 corresponds to severe apathy where the patient feels apathetic all the time^36^.

### Statistical analyses Cross-sectional analyses

The following linear regression models were used to investigate the cross-sectional relationships between brain measurements (i.e. regional DBM and WMH values) and apathy scores at baseline.

***Apathy*** *∼ Brain Measure + Age + Sex + Motion rating + TIV + HY Stage*

The variable of interest was *Brain Measure*, indicating the associations between the brain measures (i.e. regional DBM and WMHs) and apathy after accounting for covariates. Age, sex, Total Intracranial Volume (TIV)^53^, motion rating, and HY Stage were included as covariates in the models.

### Longitudinal analyses

A series of mixed-effect models were used to investigate the relationship between baseline brain measures and longitudinal changes in apathy scores. The following linear mixed-effects models were conducted to examine whether regional DBM, total, or regional WMH would influence the apathy score:

***Apathy*** *∼ Brain Measure Baseline + Years from Baseline + Brain Measure Baseline : Years from Baseline + Age Baseline + Sex + Motion rating Baseline +Apathy Baseline + TIV + LEDD + HY Stage +(1|Patient ID)*

The variable of interest was the interaction term *Brain Measure:Years from Baseline*, reflecting how baseline atrophy and WMHs contribute to future apathy in PD. Age at baseline, sex, motion rating (corresponding to the baseline MRI), HY Stage, baseline apathy scores, TIV, and LEDD were included as covariates. Patient ID was included as a categorical random effect.

### Hypothesis-based and exploratory analyses

The DBM analyses were performed at two levels: i) hypothesis-based, assessing the impact of atrophy in brain regions reported to be associated with apathy in PD in the literature^14–23^, and ii) exploratory, assessing the potential impact of atrophy in all CerebrA GM regions to determine if any new regions could be identified. Table 1 presents the list of the regions included in the hypothesis-based analyses and whether their atrophy was related to apathy presence or severity from the literature. Similar analyses were also completed for global as well as regional WMHs. All analyses (i.e. GM and WMHs) were corrected for multiple comparisons using the False Discovery Rate (FDR) controlling technique^54^. All statistical analyses were performed using MATLAB version 2022a. fitlm and fitlme functions were used to complete the linear regression and mixed effects models, respectively.

## Results

Figure 2 summarizes the results of the quality control and participant selection process. The baseline and longitudinal demographics and clinical characteristics of the PD patients included in this study are summarized in Table 2. Among all PD participants in the PPMI, our study focused on the subset of 546 individuals with idiopathic PD that had T1w MRI available at the initial visit, excluding genetic variants and SWEDD cases. A total of 445 PD patients had baseline MRI and clinical assessments available, with ages between 33.8 and 84.9. Additionally, 430 participants had longitudinal clinical assessments available, resulting in a final longitudinal dataset of 430 patients with 5068 timepoints. There were no significant differences in demographic and clinical characteristics of the included patients and those that were excluded based on MRI QC (Table S.1 in Supplementary Materials).

**Table 2.** Characteristics of the participants included in this study.

| Measure | Baseline-only Sample | Longitudinal Sample at Baseline | Longitudinal Timepoints |
| --- | --- | --- | --- |
| **N (Visit)** | **445 (445)** | **430 (430)** | **430 (5068)** |
| **Sex (%female)** | 150(33.7) | 145(33.7) | 1676 (33.07) |
| **Age** | 62.42 ± 9.62 | 62.27 ± 9.63 | 64.38 ± 9.88 |
| **Years from Baseline** | - | - | 3.21 ± 3.1 |
| **Education Years** | 15.99 ± 2.99 | 16.01 ± 2.97 | 15.88 ± 2.85 |
| **MOCA** | 27.10 ± 2.26 | 27.01 ± 2.48 | 26.75 ± 3.03 |
| **LEDD** | - | - | 364.15 ± 388.41 |
| **Apathy Score** | 0.24 ± 0.57 | 0.23 ± 0.57 | 0.35 ± 0.68 |
| **GDS** | 2.30 ± 2.48 | 2.27 ± 2.47 | 2.60 ± 2.76 |
| **HY Stage** | 1.61 ± 0.50 | 1.62 ± 0.50 | 1.87 ± 0.55 |
| **UPDRS-III** | 21.41 ± 8.96 | 21.39 ± 9.09 | 25.94 ± 12.08 |

MoCA: Montreal Cognitive Assessment. LEDD: levodopa-equivalent daily dose. GDS: Geriatric Depression Scale. HY: Hoehn and Yahr. UPDRS: Unified Parkinson's Disease Rating Scale.

### Hypothesis-based analyses

No cross-sectional regional associations survived multiple comparison corrections. The longitudinal hypothesis-based models revealed significant interaction effects between baseline atrophy (i.e., lower DBM values) and time in relation to apathy in the bilateral accumbens area, superior parietal, putamen, insula, left precuneus, right precentral, and cerebellum gray matter. (Fig. 3., Table S.2 in the supplementary materials).

**Figure 3.**
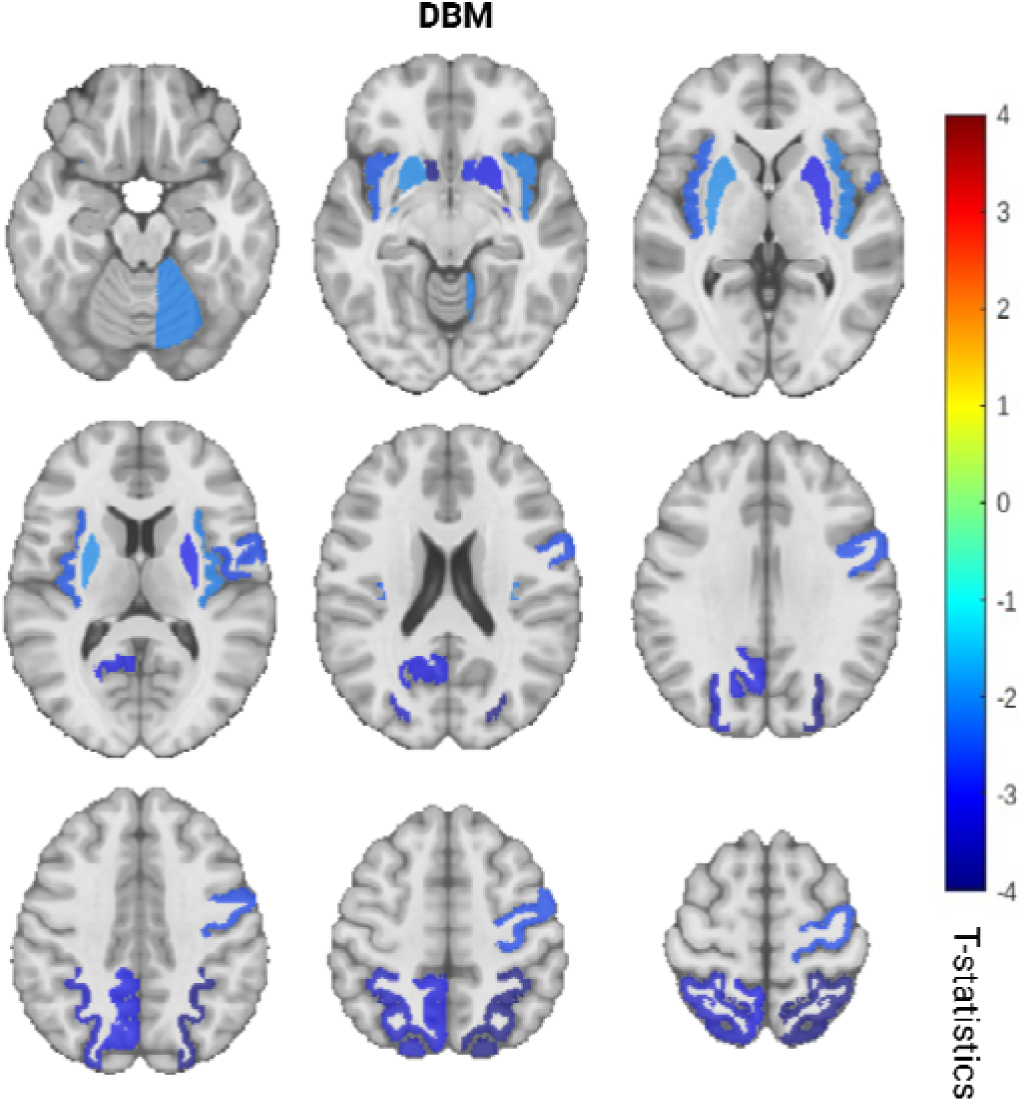
Longitudinal hypothesis-based analyses results. Cold colors indicate the t-statistics values corresponding to the significant longitudinal associations between regional atrophy and severity of future apathy.

### Exploratory analyses

No cross-sectional regional associations survived multiple comparison corrections in the exploratory analyses for either DBM or WMH measurements. Longitudinal exploratory analyse further revealed significant interactions between baseline atrophy and time impacting future apathy in the bilateral lingual, parahippocampal, basal forebrain, ventral diencephalon, isthmus cingulate, thalamus, hippocampus, the left middle temporal, and right hemisphere, including th inferior temporal, peri calcarine, medial orbitofrontal, and cuneus. These results suggest that baseline atrophy in these regions may impact the longitudinal trajectory of apathy in PD (Fig.4.A, Table S.3 in the supplementary materials). Longitudinal assessment of WMHs also showed significant interactions of bilateral frontal lobe WMH burden at baseline and tim impacting future apathy (Fig.4.B, Table S.4 in the supplementary materials).

**Figure 4.**
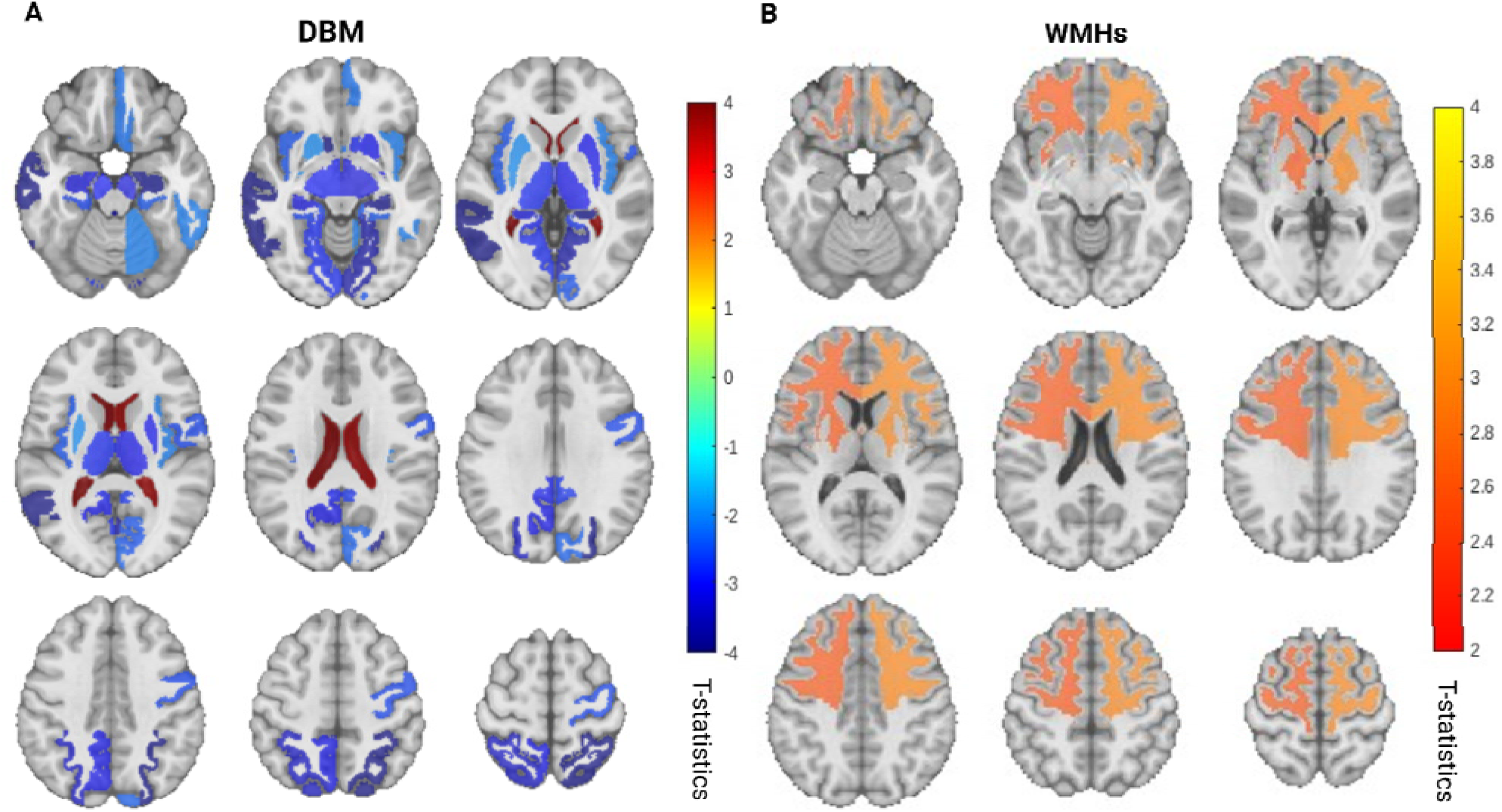
Longitudinal exploratory analysis results. A. Cold colors indicate the t-statistics values corresponding to the significant longitudinal associations between baseline regional atrophy and severity of future apathy. Warm colors indicate the significant longitudinal associations between baseline ventricular expansion and severity of future apathy. B. Colors indicate the t-statistics values corresponding to the significant longitudinal associations betwee baseline WMH burden and severity of future apathy.

## Discussion

The present study investigated the underlying neural correlates of apathy in PD by utilizing a large dataset with longitudinal apathy assessments. Our findings emphasize the importance of specific brain regions in manifestation of apathy in de novo PD. Our cross-sectional analyses did not reveal any associations between apathy severity and regional atrophy at baseline when patients were at the de novo stage. Utilizing baseline DBM data and multiple apathy measurements per participant, our longitudinal mixed-effects modeling analyses confirmed significant associations between future apathy and baseline atrophy in the bilateral accumbens area^15,17,21^, superior parietal^17^, putamen^19^, insula^14,16,19,23^, left precuneus^14^, precentral^14,16^, and right cerebellum GM^20^.

To further examine the relationship between brain structural changes and apathy severity, we conducted exploratory analyses using cross-sectional and longitudinal approaches. While we found no regions to show significant associations with apathy severity cross-sectionally, our longitudinal exploratory analysis revealed significant associations of atrophy in several region with changes in apathy severity over time. This highlights the importance of considering disease progression in understanding the neural underpinnings of apathy in PD. The increased statistical power provided by our large sample size allowed us to extend previous findings in the literature regarding the association between brain atrophy and apathy in PD. The absence of significant results in cross-sectional analyses suggests that the relationship between atrophy and apathy may be weak at baseline de novo stages and become more apparent with disease progression and therefore can be identified using a longitudinal framework. Our study revealed additional regions linked to apathy in PD, providing a more comprehensive understanding of the mechanisms underlying apathy in PD.

Associations between apathy and atrophy in regions such as the accumbens, putamen, insula, thalamus, and hippocampus suggest contributions to the motivational apathy subtype in this sample. This subtype is characterized by diminished reward sensitivity and reduced intrinsic motivation^6,11^. In addition, this interpretation is supported by atrophy in the parahippocampal, ventral diencephalon, and cingulate isthmus, implicating these regions in reward processing and emotional regulation^6,11^. It is worth noting that in areas related to functional impairment such as the medial frontal cortex, orbitofrontal cortex, and caudate nucleus, no significant relationship was found between the progression of apathy and atrophy ^6^. This suggests that cognitive apathy, associated with executive dysfunction, is less prominent in this group of PD patients^6^.

Utilizing quantitative volumetric assessments, we found that frontal lobe WMH burden at baseline is strongly associated with the increase in apathy scores over time. Our lobar WMH segmentation allows us to examine the correlation of WMH burden with apathy progression both at the whole brain level and within specific brain lobes. This approach provided a more precise understanding of brain changes associated with apathy, which was previously unexamined^30^. Our results suggest that early WMH burden at the frontal lobes is significantly associated with worsening apathy in the future. As WMHs can be managed (e.g. through antihypertensive medications), targeting vascular pathology early might offer a therapeutic avenue to mitigate the progression of apathy in PD patients^32^. This suggests the importance of incorporating WMH burden assessments in routine clinical evaluations to better predict and manage apathy in PD.

The image processing pipelines used in this study have been specifically developed and extensively validated for use in multi-center and multi-scanner applications, and have previously been employed in numerous studies to investigate atrophy and WMH burden in aging^55^ and different neurodegenerative disorders such as Alzheimer’s disease^45,56^, frontotemporal dementia^57,58^, amyotrophic lateral sclerosis^59,60^, multiple sclerosis^61^, and PD^62–65^. Further still, all image processing steps were quality controlled to ensure the accuracy of the derived atrophy and WMH measurements. Our study also has limitations that need to be taken into account. While the UPDRS-I is used in the PPMI dataset, its accuracy in assessing apathy is limited compared to other more comprehensive apathy assessment tools. In this regard, Pagonabarraga et al^11^. highlight that while the UPDRS-I is useful for screening purposes, it might lack the depth and specificity needed to assess apathy at PD. In contrast, scales such as the SAS and the LARS are more robust in assessing apathy as they investigate a more comprehensive set of dimensions of apathy, including motivation, emotional response, and goal-directed behavior. In addition, NPI is also more effective than UPDRS-I in capturing the multifaceted nature of apathy. Thus, future research should consider integrating multiple apathy assessment tools and explore the onset and progression of neuropsychiatric symptoms in PD. Furthermore, the PPMI dataset does not include FLAIR images for all participants which would be optimal for WMH assessments. While our T1w-based WMH measurements have been shown to hold strong correlations with FLAIR-based WMHs (r = 0.96, p < 0.001), future studies should also assess the relationship between apathy and FLAIR-based WMHs, which would be better indicators of the extent of WM damage^44^.

## Conclusion

Our study highlights the relationships between regional brain atrophy and WMH burden and future apathy in PD, providing important insights into the neural basis of apathy. Recognizing the contribution of WMHs to future apathy in PD provides an opportunity for timely interventions, improving the quality of life in the patients.

## Data Availability

For up-to-date information on the study, visit www.ppmi-info.org. PPMI is sponsored and partially funded by the Michael J Fox Foundation for Parkinson's Research and funding partners, including AbbVie, Avid Radiopharmaceuticals, Biogen, Bristol-Myers Squibb, Covance, GE Healthcare, Genentech, GlaxoSmithKline (GSK), Eli Lilly and Company, Lundbeck, Merck, Meso Scale Discovery (MSD), Pfizer, Piramal Imaging, Roche, Servier, and UCB (www.ppmi-info.org/fundingpartners).

## Acknowledgements

Data used in this article were obtained from the Parkinson’s Progression Markers Initiative (PPMI) database (www.ppmi-info.org/data). For up-to-date information on the study, visit www.ppmi-info.org. PPMI is sponsored and partially funded by the Michael J Fox Foundation for Parkinson’s Research and funding partners, including AbbVie, Avid Radiopharmaceuticals, Biogen, Bristol-Myers Squibb, Covance, GE Healthcare, Genentech, GlaxoSmithKline (GSK), Eli Lilly and Company, Lundbeck, Merck, Meso Scale Discovery (MSD), Pfizer, Piramal Imaging, Roche, Servier, and UCB (www.ppmi-info.org/fundingpartners). The authors also acknowledge use of Compute Canada (https://alliancecan.ca/en) resources for performing the image processing analyses in the presented work.

## Supplementary Materials

**Table S.1.** Comparison of the demographics and clinical variables between the participants included and excluded in the study.

| Variable | Excluded | Included | P Value |
| --- | --- | --- | --- |
| Age | 63.317 ± 10.093 | 62.177 ± 9.635 | 0.3 |
| Sex | 1.667 ± 0.475 | 1.661 ± 0.474 | 0.93 |
| Education_Years | 16.000 ± 3.379 | 15.979 ± 2.992 | 0.997 |
| MOCA | 26.556 ± 2.926 | 27.110 ± 2.271 | 0.296 |
| GDS | 1.789 ± 2.261 | 2.268 ± 2.462 | 0.045 |
| Apathy Score | 0.099 ± 0.300 | 0.233 ± 0.563 | 0.079 |
| HY Stage | 1.618 ± 0.490 | 1.613 ± 0.501 | 0.913 |
| UPDRS III | 22.099 ± 8.469 | 21.394 ± 9.003 | 0.361 |

**Table S.2.** Hypothesis-based results for the longitudinal DBM analyses.

| Hemisphere | Left |  |  |  | Right |  |  |  |
| --- | --- | --- | --- | --- | --- | --- | --- | --- |
| Region Names | ID | T stats | P values | P<sub>FDR</sub> | ID | T stats | P values | P<sub>FDR</sub> |
| Middle Frontal Gyrus | 52 | 0.050 | 0.960 | 0.993 | 1 | -1.004 | 0.316 | 0.451 |
| **Nucleus Accumbens** | **55** | **-4.406** | **<0.001** | **<0.001** | **4** | **-3.305** | **0.001** | **0.005** |
| Orbitofrontal Cortex | 58 | -2.101 | 0.036 | 0.082 | 7 | -2.285 | 0.022 | 0.056 |
| Rostral Anterior Cingulate | 59 | -0.880 | 0.379 | 0.516 | 8 | 0.432 | 0.666 | 0.799 |
| **Superior Parietal Gyrus** | **60** | **-3.831** | **<0.001** | **0.001** | **9** | **-4.385** | **<0.001** | **<0.001** |
| Inferior Parietal Gyrus | 61 | -1.410 | 0.159 | 0.280 | 10 | 0.351 | 0.726 | 0.838 |
| **Putamen** | **72** | **-2.374** | **0.018** | **0.048** | **21** | **-3.435** | **0.001** | **0.004** |
| **Insula** | **74** | **-2.891** | **0.004** | **0.014** | **23** | **-2.540** | **0.011** | **0.037** |
| **Precuneus** | **82** | **-3.478** | **0.001** | **0.004** | 31 | -1.513 | 0.130 | 0.245 |
| **Precentral Gyrus** | 86 | -0.098 | 0.922 | 0.988 | **35** | **-3.032** | **0.002** | **0.010** |
| Caudal Midbrain | 93 | -0.610 | 0.542 | 0.678 | 42 | -0.625 | 0.532 | 0.678 |
| Inferior Frontal Gyrus | 95 | 1.964 | 0.050 | 0.106 | 44 | 1.349 | 0.177 | 0.296 |
| Superior Temporal Gyrus | 96 | -1.250 | 0.211 | 0.317 | 45 | -1.256 | 0.209 | 0.317 |
| **cerebellum gray matter** | 97 | -1.673 | 0.094 | 0.189 | **46** | **-2.494** | **0.013** | **0.038** |
| Caudate | 100 | 0.306 | 0.759 | 0.844 | 49 | 0.008 | 0.994 | 0.994 |

**Table S.3.** Exploratory results for the longitudinal DBM analyses.

| Right Hemisphere |  |  |  | Left Hemisphere |  |  |  |
| --- | --- | --- | --- | --- | --- | --- | --- |
| Region Names | ID | T stats | P<sub>FDR</sub> | Region Names | ID | T stats | P<sub>FDR</sub> |
| Inferior temporal | 3 | -2.5139 | 0.0382 | Accumbens Area | 55 | -4.4058 | <0.001 |
| Accumbens Area | 4 | -3.3049 | 0.0051 | Superior Parietal | 60 | -3.8314 | 0.0013 |
| Pericalcarine | 6 | -2.8933 | 0.0146 | Lingual | 63 | -3.2757 | 0.0053 |
| Superior Parietal | 9 | -4.3848 | <0.001 | Para hippocampal | 69 | -3.0439 | 0.01 |
| Lingual | 12 | -4.113 | <0.001 | Putamen | 72 | -2.374 | 0.05 |
| Medial Orbitofrontal | 15 | -2.6134 | 0.0306 | Insula | 74 | -2.8908 | 0.0146 |
| Para hippocampal | 18 | -2.8694 | 0.015 | Basal Forebrain | 76 | -5.0842 | 0 |
| Putamen | 21 | -3.4351 | 0.0041 | Ventral Diencephalon | 77 | -3.3631 | 0.0047 |
| Insula | 23 | -2.5401 | 0.0366 | Middle Temporal | 79 | -4.9283 | 0 |
| Basal Forebrain | 25 | -5.3788 | 0 | Precuneus | 82 | -3.4779 | 0.0037 |
| Ventral Diencephalon | 26 | -3.6484 | 0.0025 | Isthmus Cingulate | 84 | -3.359 | 0.0047 |
| Isthmus Cingulate | 33 | -3.1874 | 0.0067 | Fourth Ventricle | 88 | -3.895 | 0.0011 |
| Precentral | 35 | -3.0322 | 0.01 | Thalamus | 91 | -3.2696 | 0.0053 |
| Fourth Ventricle | 37 | -4.001 | <0.001 | Lateral Ventricle | 92 | 3.3309 | 0.0049 |
| Thalamus | 40 | -3.478 | 0.0037 | Hippocampus | 99 | -3.5901 | 0.0028 |
| Lateral Ventricle | 41 | 3.0832 | 0.0091 |  |  |  |  |
| Cuneus | 43 | -2.788 | 0.0187 |  |  |  |  |
| Cerebellum Gray Matter | 46 | -2.4936 | 0.0392 |  |  |  |  |
| Hippocampus | 48 | -4.3271 | <0.001 |  |  |  |  |

**Table S.4.**
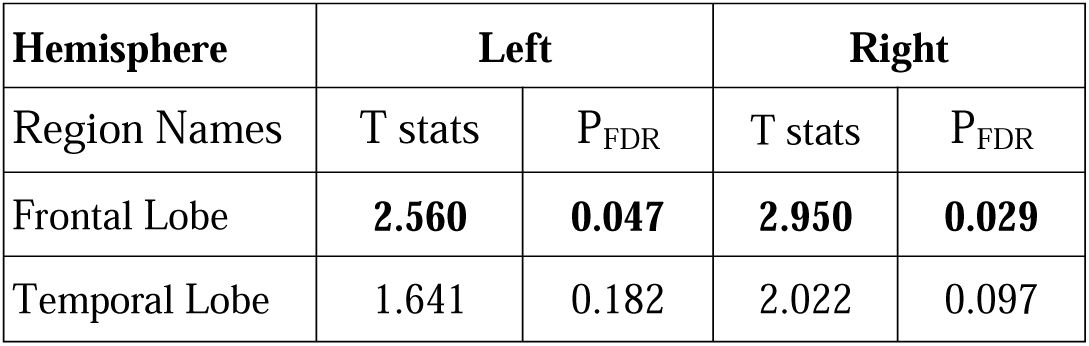
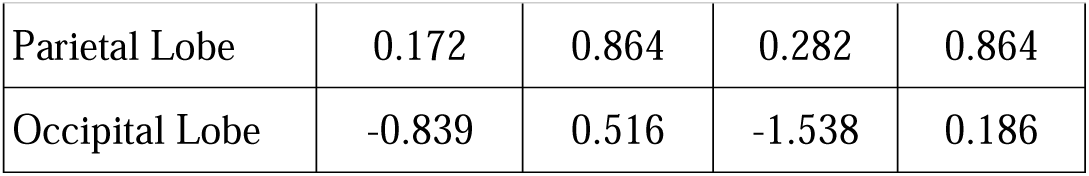
Exploratory results for the longitudinal WMH analyses.

| Hemisphere | Left |  | Right |  |
| --- | --- | --- | --- | --- |
| Region Names | T stats | P<sub>FDR</sub> | T stats | P<sub>FDR</sub> |
| Frontal Lobe | **2.560** | **0.047** | **2.950** | **0.029** |
| Temporal Lobe | 1.641 | 0.182 | 2.022 | 0.097 |
| Parietal Lobe | 0.172 | 0.864 | 0.282 | 0.864 |
| Occipital Lobe | -0.839 | 0.516 | -1.538 | 0.186 |

